# EVALUATION OF THE DURABILITY OF LONG-LASTING INSECTICIDAL NETS IN GUATEMALA

**DOI:** 10.1101/2020.07.30.20165316

**Authors:** María Eugenia Castellanos, Soledad Rodas, Jose Guillermo Juarez, Juan Carlos Lol, Sayra Chanquin, Zoraida Morales, Lucrecia Vizcaino, Stephen C. Smith, Jodi Vanden Eng, Henok G. Woldu, Audrey Lenhart, Norma Padilla

**Author notes:** Both authors contributed equally to this manuscript (María Eugenia Castellanos, Soledad Rodas). CORRESPONDING AUTHOR: Norma Padilla, PhD., Center for Health Studies, Universidad del Valle de Guatemala, Guatemala.gt.

## Abstract

**Background:** Insecticide-treated bednets (ITNs) are widely used for the prevention and control of malaria. In Guatemala, since 2006, ITNs have been distributed free of charge in the highest risk malaria-endemic areas and constitute one of the primary vector control measures in the country. Despite relying on ITNs for almost 15 years, there is a lack of data to inform the timely replacement of ITNs whose effectiveness becomes diminished by routine use.

**Methods:** We assessed the survivorship, physical integrity, insecticide content and bio-efficacy of ITNs through cross-sectional surveys conducted at 18, 24 and 32 months after a 2012 distribution of PermaNet® 2.0 in a malaria focus in Guatemala. A total of 988 ITNs were analyzed (290 at 18 months, 349 at 24 months and 349 at 32 months).

**Results:** The functional survivorship of bednets decreased over time, from 92% at 18 months, to 81% at 24 months and 69% at 32 months. Independent of the time of the survey, less than 80% of the bednets that were still present in the household were reported to have been used the night before. Most of the bednets had been washed at least once (88% at 18 months, 92% at 24 months and 96% at 32 months). The proportion of bednets categorized as “in good condition” per WHO guidelines of the total hole surface area, diminished from 77% at 18 months to 58% at 32 months. The portion of ITNs with deltamethrin concentration less than 10mg/m^2^ increased over time (14% at 18 months, 23% at 24 months, and 35% at 32 months). Among the bednets for which bioassays were conducted, the percentage that met WHO criteria for efficacy dropped from 90% at 18 months to 52% at 32 months.

**Conclusion:** While our assessment demonstrated that nets were in relatively good physical condition over time, the combination of declining bio-efficacy over time and low use rates limited the overall effectiveness of the LLINs. Efforts to encourage the community to retain, use, and properly care for the LLINs may improve their impact. Durability assessments should be included in future campaigns.

## INTRODUCTION

Globally, insecticide-treated bednets (ITNs) are a standard intervention for the prevention and control of malaria. Thirteen of the seventeen malaria-endemic countries in the Americas have incorporated within their malaria control and prevention programs the universal distribution of ITNs, and report having a policy of distributing ITNs through mass campaigns (World Health Organization, 2018).

The use of ITNs has had a significant impact on personal protection and in reducing malaria transmission. If high ITN coverage is sustained (> 80%), nets can provide protection to the larger community, including those households that do not use them (Hawley et al., 2003). The Global Malaria Program of the World Health Organization (WHO) recommends universal access and use of ITNs as a primary intervention for malaria control (World Health Organization, 2017).

Previous assessments of ITNs show a high degree of variation in ITN durability (Fettene, Balkew, & Gimblet, 2009; Kilian et al., 2011; Rosas-Aguirre et al., 2011), with ITNs from the same manufacturer showing significantly different levels of durability. The factors that influence this variability include the frequency of washing (including the quality of the water), type of laundry soap, washing and drying techniques, and daily levels of “wear and tear”. Translating this variability into a single 3-5 year durability estimate for ITNs could result in an inaccurate presumption of ITN effectiveness in areas where durability is compromised considerably faster. As such, understanding durability is a key consideration for the timing of ITN replacement strategies, and should be a component of any distribution campaign.

While ITN durability has been much more broadly studied in Africa, limited data are available from the Americas. An ITN distribution took place in Guatemala in 2012-2013 as part of malaria control activities supported by the Global Fund. Approximately 929,000 PermaNet® 2.0 long-lasting insecticidal nets (LLINs) were distributed in 14 Health Areas that comprise a total of 111 municipalities in 12 departments of Guatemala. The primary objective of this study was to assess the durability of the PermaNet^®^ 2.0 LLINs over 32 months of routine use in an area with high malaria burden in the Mesoamerican sub-region. The three elements of durability, survivorship, physical integrity and insecticidal activity were evaluated, following the WHO guidelines for monitoring the durability of ITNs under operational conditions (World Health Organization., 2011). This evaluation was designed to provide relevant data for future plans around ITN replacement strategies that could be relevant in both Guatemala and more broadly in Latin America.

## METHODS

### Study site and description of the ITN distribution

The study was conducted in the municipality of La Gomera in the department of Escuintla in southern Guatemala (Figure 1). Escuintla was included in the 2012-2013 ITN mass distribution campaign. According to national surveillance data, 56% of all malaria cases in Guatemala are reported from Escuintla (2012). As of 2012, La Gomera is divided into 19 communities, had a population of 60,299 inhabitants and reported 23% of the malaria cases in the country (PAHO, 2017). In Escuintla, malaria is mainly transmitted by *Anopheles albimanus* and *Plasmodium vivax* is responsible for over 95% of the cases. 69,214 PermaNet® 2.0 (Vestergaard Frandsen, Lausanne, Switzerland) deltamethrin-treated LLINs were distributed in Escuintla from April 2012 to September 2013. It was the first mass LLIN distribution campaign in the region.

**Figure 1.**
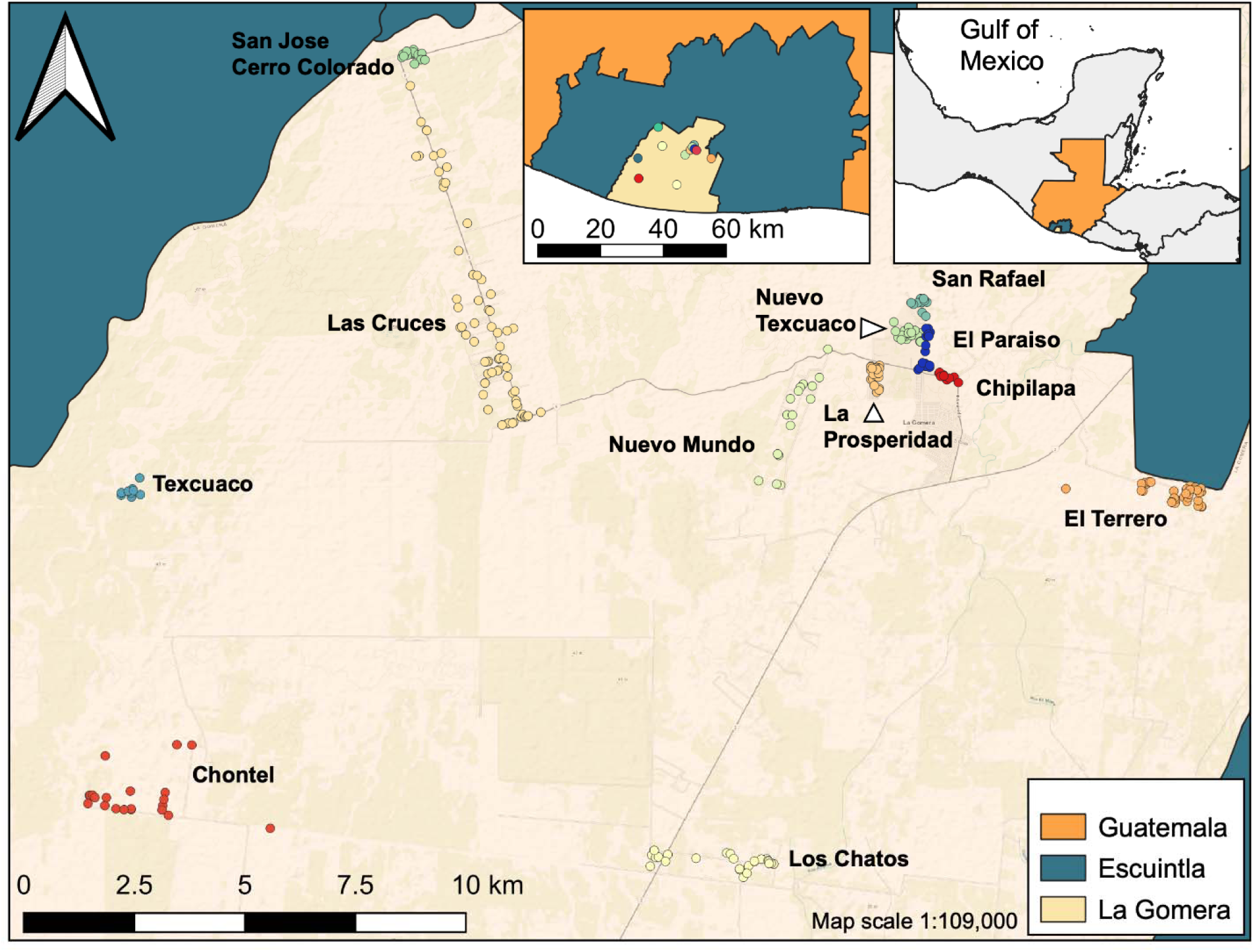
Location of the 12 study communities in La Gomera, Escuintla, Guatemala.

### Experimental design

The evaluation was carried out from November 2013 to March 2015 through three prospective cross-sectional surveys in randomly selected households using a multistage sample design. The first stage of the sampling consisted of a non-probabilistic sampling during which one municipality within the LLINs distribution area was selected (La Gomera). The following stage was done using a cluster survey methodology with each cluster size proportional to its population. The evaluation included only those communities that received LLINs approximately 18 months (range=17-19 months) prior to the first survey. Twelve communities met this criterion and were included in the 18-and 24-months survey. In this twelve communities, 4,076 had been delivered in the study area between April-June 2012. At the 32-month survey, only 8 of these 12 communities were included since in the other four communities a round of LLIN replacement occurred in 2014. The selection of households to be sampled within each stratum was done through a simple random sampling. Nets were not labelled prior to distribution and therefore users were not aware that the nets were to be evaluated, reducing the possibility of the Hawthorne effect (Earl-Slater, 2002).

### Sample size for the cross-sectional surveys

To determine survivorship and physical integrity of the LLINs, sample size estimates were based on the confidence interval around the attrition rate at each time period, according to values reported for Kenya (11% for 18 months, 15% for 24 months, 23% for 30 months) (Gimnig, 2013). We assumed a relative error of 5%, a confidence level of 95%, and a design effect of 2.0. We assumed a non-response proportion of 15% at 18 months. For the surveys conducted at 24 months and 32 months we had to increase this proportion to 40% and 55%, respectively as when we visited the households many of them reported that they had not received any LLIN. Also, for the surveys at 24 months and 32 months, we increased by 5% the number of households to survey because we had used Google Maps to select the households (more details in the next section) and it was possible that some structures that had been enumerated as households were another type of structure (Wampler, Rediske, & Molla, 2013). Thus, the numbers of households selected to survey at each time point were as follows: n=348 at 18 months, n=583 at 24 months, and n=887 at 32 months, for a total of 1,818 households.

### Selection of households and nets evaluated

For the 18-month survey, households were identified using the Ministry of Health (MoH) 2012 LLINs campaign distribution list. We used the OpenEpi Random Program (www.openepi.com) to generate a random number list for the final selection of households. We then used the addresses of the households provided by the MoH to locate the household. In the 24-month and 32-month surveys, we used aerial Google Earth imagery to identify houses for sampling (Map data: Google, DigitalGlobe). The high resolution of the 2013 images of the selected communities allowed for good accuracy when mapping individual houses. The Google Earth file was imported to ArcMap 9.3 (ESRI, Redlands, CA, USA) using a KML to shapefile extension (Parent, 2009). Longitude and latitude coordinates (WGS84 coordinate systems) were added to the attributes table which was saved as a dBASE file. This was imported to an Excel spreadsheet for the random household selection, using simple random sampling with the OpenEpi Random Program. The geographic information system (GIS) information about each selected community was uploaded to GPS Visualizer (www.gpsvisualizer.com), and subsequently added to a Garmin global positioning system (GPS) map 62S (Garmin, USA) with a 20m proximity alarm. A field validation of the maps and GPS equipment was carried out prior to conducting the fieldwork.

### Questionnaires

Program staff from the National Malaria Sub-program of the Vector-borne Diseases Program of the MoH and researchers from Universidad del Valle de Guatemala conducted the surveys. The survey teams were trained to understand the structure of the survey and how to deliver it using the personal digital assistants (PDAs) that included a GPS element. Survey questions were based on WHO Durability guidelines (World Health Organization., 2011), adapted to local settings, translated to Spanish and validated. The questionnaire was administered primarily to the adult woman in charge of the household in order to collect as much data as possible on the maintenance and washing frequencies of the LLINs. If an adult woman was not present, then the questionnaire was administered to any adult household member. Questions were included on a) demographic characteristics of the enrolled household; b) net survivorship, attrition and use; and c) physical integrity of the fabric measured *in situ*. Of the total number of LLINs that were delivered to the household, one was randomly selected for further analysis (1 enrolled household = 1 enrolled LLIN). Questions on net use and physical integrity were directed towards this randomly selected specific net if it was still present in the household. If the selected LLIN was no longer present, other questions related to reasons for its absence were pursued. In the 24-month and 32-month surveys, if a previously surveyed household was selected, we still randomly selected a LLIN to evaluate. If the selected LLIN had already been evaluated, we would select another one if available.

### Physical integrity

LLINs were assessed *in situ* with the net left hanging over the sleeping space if the position allowed it. Holes were classified according to the WHO 2011 categories: (1) holes smaller than a thumb (∼0.5 to 2 cm diameter), (2) bigger than a thumb but smaller than a fist (∼2 to 10 cm diameter), (3) bigger than a fist but smaller than a head (∼10 to 25 cm diameter) and (4) bigger than a head (> ∼ 25 cm diameter) (World Health Organization., 2011). A standard ruler was available in case there was uncertainty on a measurement.

### Insecticide content

Deltamethrin content, in mg/m^2^, was measured using a Tracer III-SD handheld X-ray fluorescence (XRF) analyzer (Bruker Nano Analytics, Inc., Kennewick, WA, USA). For this analysis, deltamethrin content was calculated from the intensity of 11.549-12.248 keV X-rays emitted by the bromine atoms of the deltamethrin in the sample (Smith et al., 2007). Calibration samples of polyester mosquito netting treated with six different levels of deltamethrin (0-114 mg/m^2^) were used to correlate intensity with deltamethrin content. The correlation was linear with R^2^=0.98, minimum. For whole bednet analysis, the net was folded in a way to create a 24-layer sampling point, comprising 8 points from the roof of the net, and 4 points from each of the 4 sides. This multilayer sample was then analyzed to yield the average deltamethrin content over the 24 locations. Since folding the net and collecting the data takes only 2-3 minutes, the average deltamethrin content of many nets can be measured quickly and conveniently (Anshebo et al., 2014; Craig et al., 2015).

### Sample size for cone bioassays

From the LLINs evaluated in the households in each community, a sub-sample of the nets reported to have been used to sleep at least once was randomly selected and collected for bio-efficacy testing. In order to detect the loss of at least 5.6% of insecticide, with respect to the baseline level of 55 mg/m^2^, a power of 90%, alpha=0.05, and using a standard deviation of 5, a subset of 55 LLINs were randomly selected during the 18-month survey, 60 at 24 months, and 65 at 32 months, with increasing number of nets selected in order to compensate for an estimated non-response effect of 15%. Collected nets were placed individually in plastic bags and grouped inside large black polyethylene bags and kept at ± 20°C until their arrival to the laboratory. Householders were given a new PermaNet^®^ 2.0 bednet to replace the collected net.

### WHO cone bioassays

Cone bioassays were performed on samples from each of the five sides of each bednet and following the sampling scheme recommended by WHO (World Health Organization., 2011). Until processed the net samples were stored at 4°C wrapped in aluminum foil and placed inside plastic bags. Briefly, we used 2-3-day old, non-blood fed *Anopheles albimanus* females from the laboratory insecticide susceptible SANARATE strain. Two replicates of 20 mosquitoes were exposed to two study PermaNet^®^ net pieces and two replicates of 10 mosquitoes were exposed to an untreated polyester net as control. Mosquitoes were exposed for 3 minutes and the 60 minutes knock-down and 24-hour mortality recorded according to WHO criteria. Data were adjusted according to Abbot’s formula. Bioassays were performed under laboratory conditions, 27 ± 2 °C and 70 ± 10% relative humidity. Knock down at 60 minutes (KD60) and mortality at 24 hours were calculated as the number of mosquitoes knocked down at 60 min and the number of dead mosquitoes at 24 hours proportional to the total number exposed. Results were considered valid if the control mortality was lower than 5%; between 5-20% control mortality, results were corrected using Abbott’s formula and if control mortality was greater than 20%, the bioassays for that net were repeated (World Health Organization., 2011).

### Verification of baseline LLIN insecticidal content

From the PermaNet^®^ 2.0 batch distributed in 2012, 11 nets were retained and analyzed using XRF and WHO cone bioassays at 24 months post-distribution to assess the insecticidal content of the unused LLINs (considered time-point 0).

### Data analysis

The main outcomes of the study were overall and functional survivorship, physical integrity, insecticide content measured by XRF and bio-efficacy measured by the WHO cone bioassay.

Descriptive demographic characteristics of the households that were enrolled and details of their reported use and handling of the LLINs at the three-survey time-points (18 months, 24 months and 32 months) were estimated using proportions and measures of central tendency. The overall survivorship at each time-point was defined as the number of enrolled LLINs still present in the households divided by the total number of enrolled LLINs. The functional survivorship was calculated at each time-point as follows: number of enrolled LLINs still present and “serviceable” (see definition below) in the households divided by the number of LLINs still present + LLINs not present owed to attrition reasons (damaged and thrown away or LLIN used for other purposes) (Committee & Secretariat, 2013; Tan et al., 2016; WHO Malaria Policy Advisory Committee, 2013). We then calculated the median survival time, defined as “the time point at which the estimate of functional LLIN survival crosses the 50% mark” (WHO Malaria Policy Advisory Committee, 2013) using the following formula: 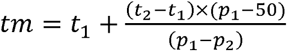 where *tm* is the median survival time, t1 and t2 were the 24 and 32 month survey points (in years) and p1 and p2 were the functional survivorship proportions for the 24 and 32 month points, respectively.

The physical integrity of LLINs was measured by estimating the proportion of LLINs with at least one hole of any size, the total number of holes/LLIN, and the total hole area (cm^2^)/ LLIN in quartiles. The total hole surface area (THSA) was calculated by multiplying the number of holes by the area of each hole (1.2, 28.3, 240.5 or 706.9 depending on the size category of the holes) and then summing across the categories (Vanden Eng et al., 2015; World Health Organization, 2013). Holes smaller than 0.5 cm were not included in the assessment. We classified the LLIN condition as good (<79 cm^2^ THSA), damaged (79-788 cm^2^ THSA) or severely torn (>=789 cm^2^ THSA) following the categorization recommended by WHO, and an LLIN was considered “serviceable” if it was classified as good or damaged (WHO Malaria Policy Advisory Committee, 2013). The proportion of LLINs with evidence of repairs was also calculated.

The median concentration and interquartile range (IQR) of deltamethrin (mg/m^2^) obtained through the XRF analyses was estimated at each time point, including unused nets. We calculated the proportions of LLINs with a concentration of less than 10 mg/m^2^ and 25 mg/m^2^. These values were chosen because LLINs with less than 10 mg/m^2^ are considered to not have the minimum effective concentration of insecticide, and nets with less than 25 mg/m^2^ are considered to not contain an optimum insecticide concentration (Ashebo et al 2014).

For the bio-efficacy results, we estimated the distribution and geometric mean (CI95) for the KD60 and mortality at 24 hours, for each survey time point including unused nets (Tan et al., 2016). We also calculated the proportion (and CI95) of LLINs that showed ≥80% mortality at 24 hours, or ≥95% KD60 at each time point, including for unused nets (World Health Organization, 2013).

We assessed the association between the cone bioassay results and the deltamethrin concentration obtained by the XRF using Locally Weighted Scatterplot Smoothing (LOESS) regression analysis to estimate Kendall’s correlation coefficient. We then developed a predictive model using piecewise (segmented) regression in which we explored if the use of the deltamethrin concentration measured by XRF and adjusted by other variables (More details in Appendix) can predict if results of a bioassay on the LLIN will exceed or not the WHO threshold of ≥80% mortality at 24 hours.

We analyzed nets based on a series of sequential criteria: a) Distributed in the household, b) Still present in the household, c) Used at least once, d) Used the night before the survey, e) In serviceable condition and e) With a concentration of deltamethrin above 10 mg/m^2^ as measured by XRF (we didn’t include this criterion in the 18-month survey as we only analyzed a small number of LLINs with XRF). For the 24- and 32-month surveys, we developed a working definition of ‘LLIN providing adequate protection’, for nets that met all of these criteria. For this purpose, we estimated the proportion of LLINs that sequentially meet each criterion. In order to be analyzed in each criterion, the LLINs should have met the requirement of the previous criterion. We then estimated the overall proportion of LLINs providing adequate protection based on our study definition.

We used the Cochran-Armitage Trend Test to assess trends across the time-points and the different categorical outcomes. For continuous outcomes, we used the Kruskal-Wallis test (differences among all groups) and the Wilcoxon rank sum test (pairwise comparisons).

All analyses were conducted using SAS software v 9.4 (SAS Institute, Cary, NC, US) and R v3.6.0 (R Foundation for Statistical Computing, Vienna, Austria, 2016).

## RESULTS

### Descriptive characteristics of the study population

A total of 1,013 LLINs were enrolled in the study, of which 988 (98%) of them were considered to have valid information. Of these, 290 were evaluated at 18 months, 349 at 24 months and 349 at 32 months (Figure 2). The education level of the head of household and the proportion of households with electricity that participated in the surveys remained consistent throughout the duration of the study (Table 1). We observed a two-fold increment in the proportion of households that owned a flush toilet and that had piped water as their source of drinking water in the 32-month survey as compared with the previous time points. The median number of individuals that slept in the household the night before the survey was four (Supplemental material, Table S1). According to the census of LLINs distributed, each household received a median of three LLINs, which corresponded to the median number of sleeping places used in the household the night before of the survey.

**Figure 2.**
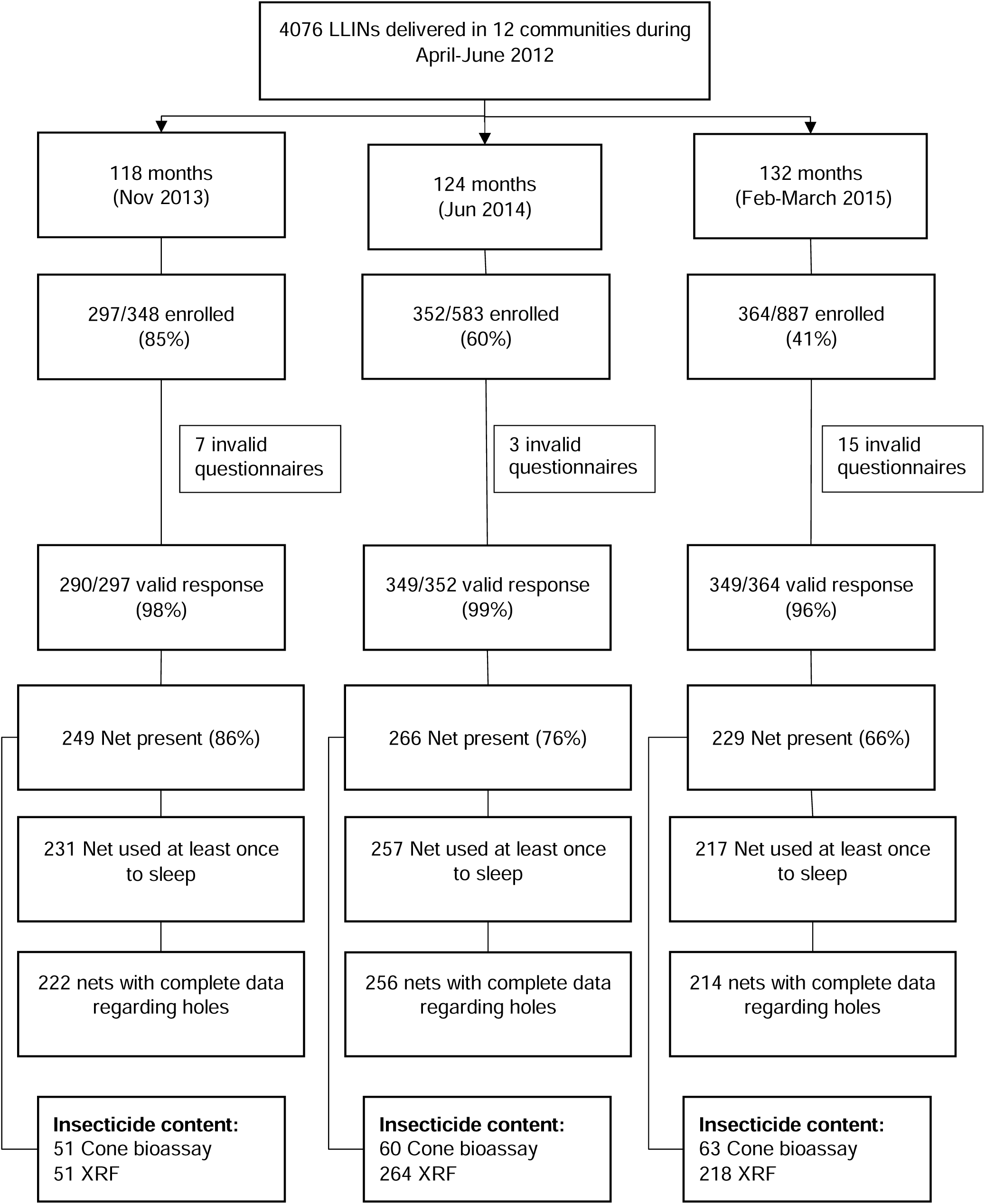
Flow chart of the data collection and number of long-lasting insecticide-treated bednets (LLINs) that were evaluated for each of the measurements. Invalid questionnaires represent questionnaires with incomplete or discrepant data or questionnaires carried out on nets that were not part of the PermaNet 2.0 2012 distribution.

### Overall and functional survivorship

The proportion of LLINs still present in the household decreased over time (p <0.001), with an overall survivorship of 86% (CI95: 82-90) at 18 months, 76% (CI95: 72-81) at 24 months and 66% (CI95: 61-71) at 32 months. The main reported cause of loss at 18 months (56%, CI95: 40-72) and 24 months (54%, CI95: 43-65) was that the net was given away to others, whereas at the 32-month survey, the main cause was that the net was damaged and thrown away (43% CI95: 34-52) (Figure 3 and Table S2). The functional survivorship was 92% (CI95: 88-95) at 18 months, 81% (CI95: 76-86) at 24 months and 69% (CI95: 64-75) at 32 months. The median survival time was extrapolated to be 3.7 years (CI95: 3.4-4.2).

**Figure 3.**
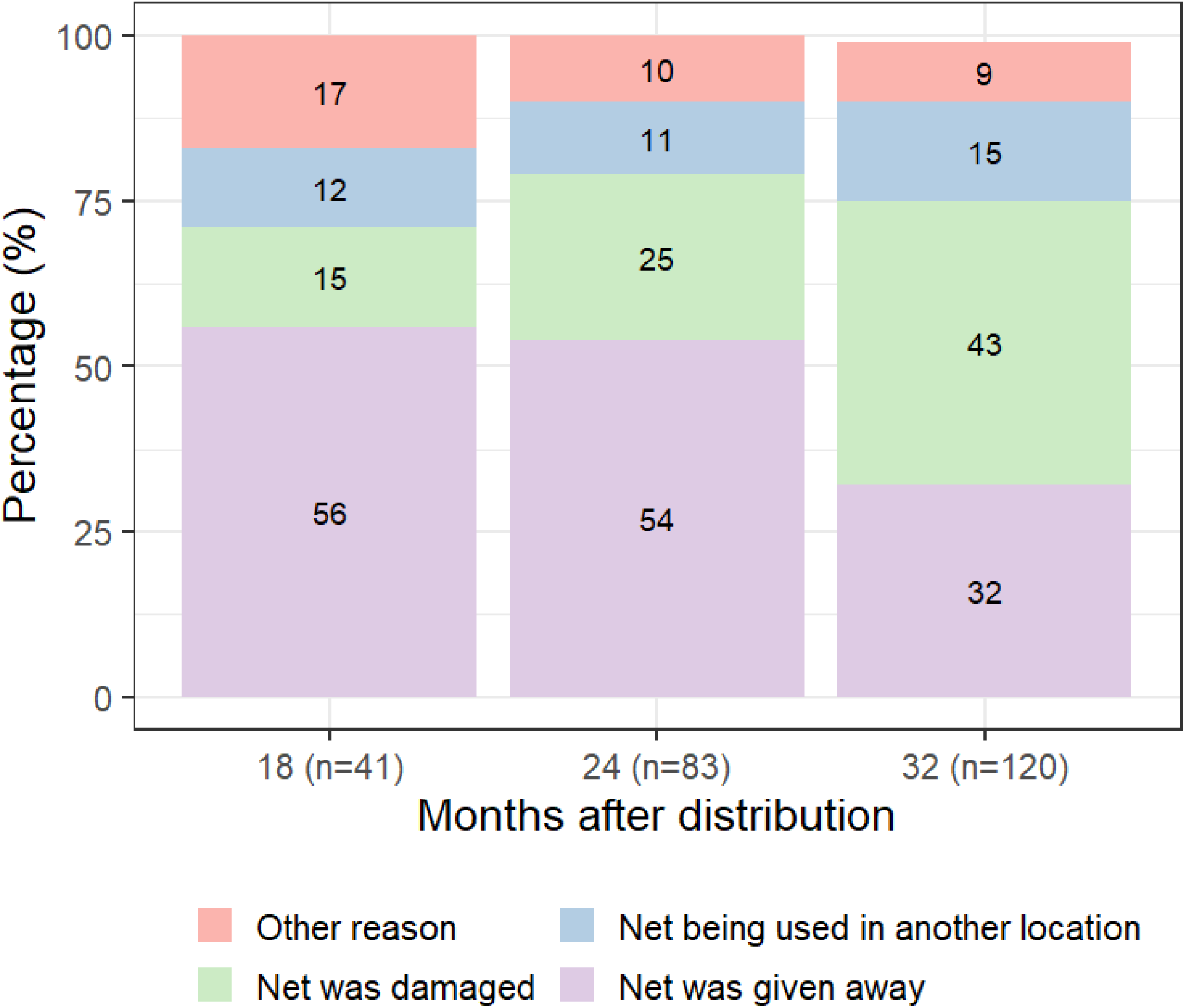
Reasons for long-lasting insecticide-treated bednet (LLIN) loss.

### LLIN use and handling

More than 90% of the LLINs still present in the households were reported to have been used at least once at any of the time points (93% at 18 months, 97% at 24 months and 95% at 32 months). However, among the LLINs used at least once, less than 80% were reported to have been used the night before the survey (74% at 18 months, 79% at 24 months and 71% at 32 months) (Table 2). Almost a third of these LLINs were found not hanging (stored away or visible but not hung up) at each of the survey time-points.

A large proportion of the LLINs that were used at least once had been washed (88% at 18 months, 92% at 24 months, 96% at 32 months) and in >55% of the cases, nets had been washed less than 1 month preceding the survey (Table 2). Among the washed LLINs, the majority were soaked (64% at 18 months, 62% at 24 months, 60% at 32 months), although in over 70% of these, the reported soaking time was less than one hour (80% at 18 months, 73% at 24 months, 74% at 32 months). The principal soap used to wash the LLINs was detergent powder, either alone or in combination with bar soap and/or bleach**Error! Reference source not found**.. Over time, there was a reduction in the proportion of LLINs that were scrubbed hard or beaten on a hard surface during washing (62% at 18 months, 51% at 24 months and 46% at 32 months, p =0.0007). In most cases, the washed LLINs were dried outside in the sun (70% at 18 months, 69% at 24 months and 60% at 32 months).

### Physical integrity

The median number of holes in the LLINs was 2 (IQR 0,7) at 18 months, 4 (0,15) at 24 months and 5.5 (IQR 1,16) at 32 months (Table 3). The median total hole area (cm^2^) in the bednets was 2.4 (IQR 0,63.8) at 18 months, 8.4 (IQR 0-271.5) at 24 months and 34.3 (IQR 1.2,327.8) at 32 months. The proportion of LLINs considered in good condition decreased from 77% at 18 months to 70% at 24 months and to 58% at 32 months (p<0.0001). At 32 months, 84% of the LLINs were considered still in ‘serviceable’ condition.

**Table 3.**
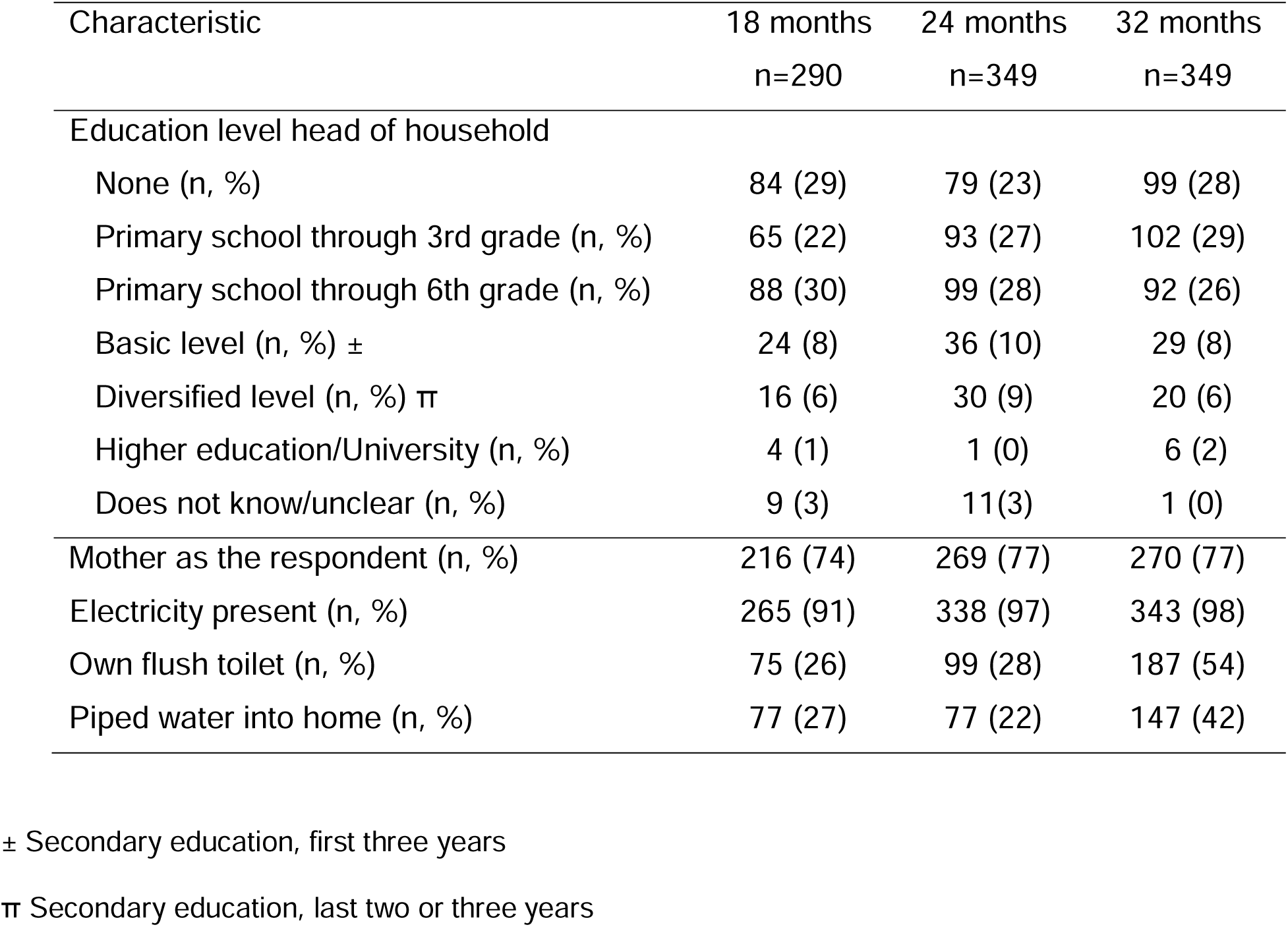
Demographic characteristics of the surveyed households and head of household at the three time-points.

Respondents reported new holes in the month prior to the survey in 24% of bednets at 18 months, in 15% of bednets at 24 months and in 27% at 32 months. The principal reported cause of these new holes was that the bednet tore or was split when caught on an object (45% at 18 months, 31% at 24 months and 38% at 32 months). Other reported causes of holes were children (21% at 18 months, 10% at 24 months and 19% at 32 months) and animals (11% at 18 months, 13% at 24 months and 10% at 32 months). However, a large proportion of these new holes did not have a known cause reported by the respondent (21% at 18 months, 36% at 24 months and 24% at 32 months). No new holes were reported due to burns. A minority of the LLINs with holes had been repaired (23% at 18 and 24 months, 44% at 32 months), with stitches as the principal mode of repair (Table 3).

**Table 3.** Physical integrity of long-lasting insecticide-treated bednets (LLINs), used at least once for sleeping and with complete data regarding holes.

| Characteristic | 18 months<br>n=222 | 24 months<br>n=256 | 32 months<br>n=214 |
| --- | --- | --- | --- |
| Any holes (n, %) | 131 (59) | 181 (71) | 162 (76) |
| Any holes repaired (n, %)± | 30 (23) | 42 (23) | 71 (44) |
| By stitches (n, %) | 23 (77) | 27 (64) | 54 (76) |
| By knots (n, %) | 9 (30) | 20(48) | 33 (46) |
| By patches (n, %) | 1 (3) | 0 (0) | 1 (1) |
| Median total number of holes [IQR] | 2 [0,7] | 4 [0,15] | 5.5 [1,16] |
| Median total hole area (cm<sup>2</sup>) [IQR] | 2.4 [0,63.8] | 8.4 [0,271.5] | 34.3 [1.2,327.8] |
| Categorization based on total hole area (cm<sup>2</sup>) |  |  |  |
| *Serviceable condition*<sup>††</sup> (n, %) | 208 (94) | 225 (88) | 180 (84) |
| Good condition (n, %) | 171 (77) | 180 (70) | 125 (58) |
| Damaged condition (n, %) | 37 (17) | 45 (18) | 55 (26) |
| *Too torn condition* (n, %) | 14 (6) | 31 (12) | 34 (16) |

Abbreviations: IQR = Interquartile boundary values (p25, p75)
± This 'n' serves as the denominator for the type of repair categories.
<sup>††</sup>Serviceable condition includes long-lasting insecticide-treated bednets with good or damaged condition.

**Table 4.** Long-lasting insecticide-treated bednets (LLINs) use and handling as reported by householders at each survey time-point.

|  | 18<br>months<br>n=249 | 24<br>months<br>n=266 | 32<br>months<br>n=229 |
| --- | --- | --- | --- |
| LLIN used at least once to sleep (n, %) ± | 231 (93) | 257 (96) | 217 (95) |
| LLIN used the night before the survey (n, %) | 171 (74) | 203 (79) | 154 (71) |
| LLIN used every night during the week before the survey (n, %) | 189 (82) | 208 (81) | 167 (77) |
| LLIN used year-round (n, %) | 205 (89) | 215 (84) | 178 (82) |
| LLIN tucked under bed at night (n, %) | 213 (92) | 241 (94) | 194 (89) |
| LLIN observed at time of survey, among LLIN used at least once to sleep |  |  |  |
| Hanging loose over sleeping place (n, %) | 66 (29) | 87 (34) | 69 (32) |
| Hanging tied in knot (n, %) | 17 (7) | 33 (13) | 35 (16) |
| Hanging folded (n, %) | 37 (16) | 28 (11) | 24 (11) |
| Visible but not hanging (n, %) | 13 (6) | 15 (6) | 10 (5) |
| Stored away (n, %) | 53 (23) | 56 (22) | 60 (28) |
| Hanging next to the wall (n, %) | 45 (19) | 38 (15) | 19 (9) |
| Washing activities, among LLIN used at least once to sleep |  |  |  |
| Ever washed <sup>π</sup> (n, %) | 204 (88) | 237 (92) | 209 (96) |
| Bed net washed <1 month ago (n, %) | 128 (63) | 131 (55) | 121(58) |
| When washed, soaked (n, %) | 130 (64) | 147 (62) | 126 (60) |
| Detergent used (n, %) | 148 (72) | 189 (80) | 163 (78) |
| When washed, scrubbed hard (n, %) | 127(62) | 122 (51) | 97(46) |
| Dried outside in the sun (n, %) | 143 (70) | 163 (69) | 126 (60) |
| Dried outside in the shade (n, %) | 54 (26) | 70 (30) | 75 (36) |
| Dried inside (n, %) | 5 (2) | 3 (2) | 4 (2) |

Abbreviations: LLIN= long-lasting insecticide-treated bednets, IQR = Interquartile boundary values (p25, p75)
± This 'n' serves as the denominator for the subsequent categories of use and position of LLIN observed at time of survey.
π This 'n' serves as the denominator for the subsequent washing categories.

### Insecticide content

The median deltamethrin concentrations (mg/m^2^) measured by XRF differed significantly by survey time-point (p<0.0001). For unused nets, the median was 49.8 mg/m^2^ (IQR 47.2, 53.0, n=11), at 18 months it was 23.2 mg/m^2^ (IQR 11.2,31.2, n=51), at 24 months it was 14.6 mg/m^2^ (IQR 8.8, 25.6, n=264) and at 32 months it was 13.9 mg/m^2^ (IQR 6.6,30.0, n=218). The reductions in insecticide content were statistically significant between unused nets and values measured at 18 months (p<0.001) and between 24 months and 32 months (p=0.027), but not between18 months to 24 months (p=0.21) (Figure 4). The proportion of LLINs with a deltamethrin concentration of less than 10 mg/m^2^ increased over time: 0% for unused nets, 14% (CI95: 4-24) at 18 months, 23% (CI95: 18-29) at 24 months and 35% (CI95: 29-42) at 32 months (p<0.0001). When the cut-off was set at 25 mg/m^2^, the proportion of LLINs below the threshold was: 0% for unused nets, 57% (CI95: 43-71) at 18 months, 69% (CI95: 63-75) at 24 months and 73% (CI95: 67-79) at 32 months (p<0.0001).

**Figure 4.**
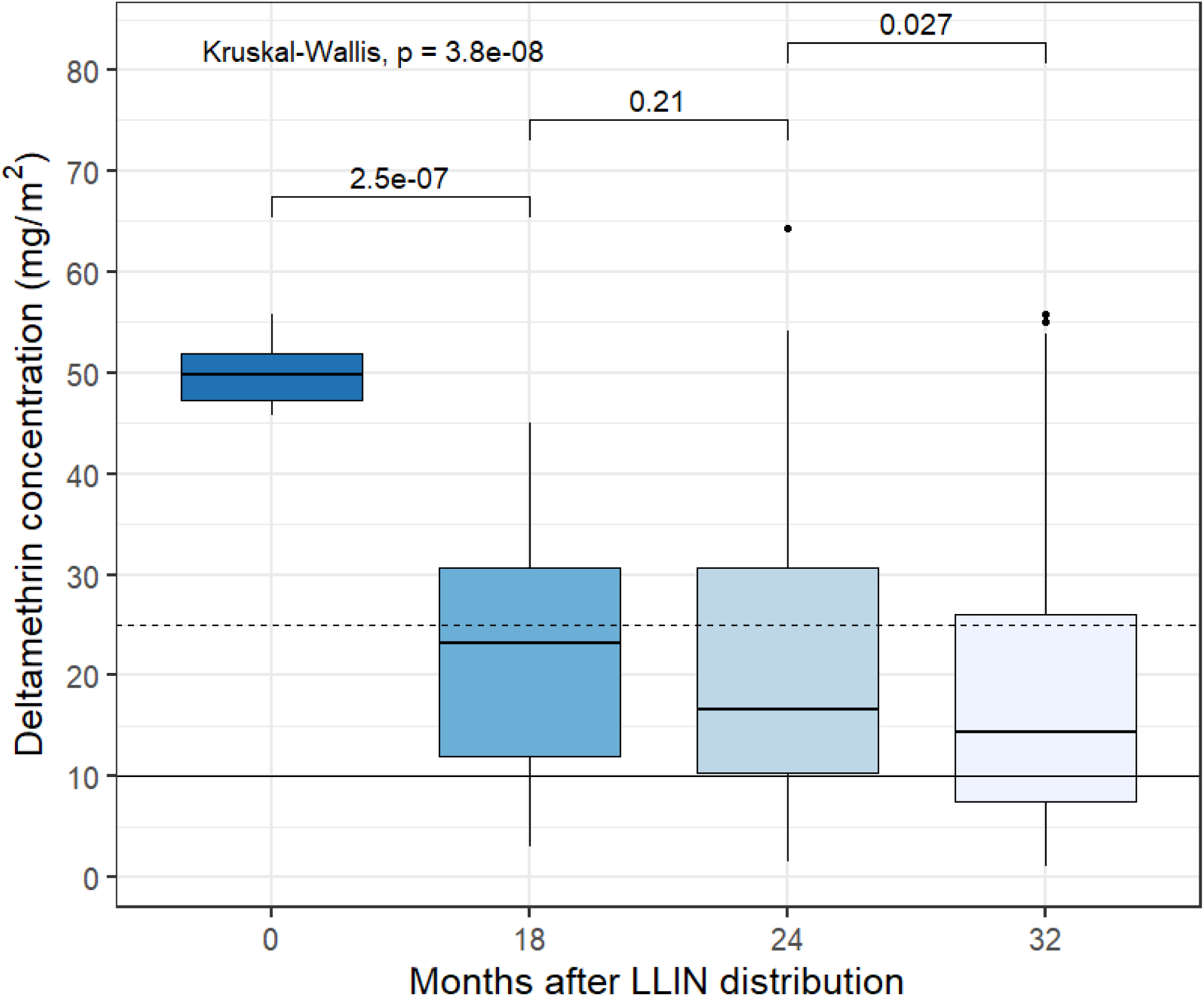
Estimated total deltamethrin concentration (mg/m^2^) measured by X-ray fluorescence (XRF) at the survey time-points. The time-point of 0 represents values on unused long-lasting insecticide-treated bednets (LLINs) that were from the same batch as those distributed. The dashed horizontal line represents the threshold of 25 mg/m^2^ and the solid horizontal line represents the threshold of 10 mg/m^2^.

### Bio-efficacy

Among the sub-set of LLINs on which bioassays were conducted, the geometric mean mortality at 24 hours decreased over time: 100% (CI95: 100,100) for unused nets (n=11), 82% (CI95: 76-89) at 18 months (n=51), 52% (CI95: 43-61) at 24 months (n=60) and 45% (CI95: 35-55) at 32 months (n=63) (Figure S1). In the case of KD60, the geometric means were 100% (CI95: 99-100) for unused nets (n=11), 96% (CI95: 94-99) at 18 months (n=51), 84% (CI95: 77-92) at 24 months (n=60) and 80% (CI95: 73-87) at 32 months (n=63) (Figure S2). On summary, the proportion of LLINs that passed the WHO≥ 80% mortality threshold or ≥95% KD60 for bio-efficacy were 100% for unused nets, 90% (CI95: 82-99) at 18 months, 68% (CI95: 56-80) at 24 months and 52% (CI95: 40-65) at 32 months (p<0.0001).

### Association between mortality at 24 hours and deltamethrin concentrations (mg/m^2^) measured by XRF

There was a positive linear relationship between deltamethrin concentration measured by XRF and the percent mortality at 24 hours up to approximately 25 mg/m^2^ of deltamethrin concentration (r=0.62, p < 0.0001) (Figure 5 & Figure S3). The summary statistics from the fitted segmented regression model confirmed the positive relationship between deltamethrin content and mosquito mortality (Table S3), with an estimated breakpoint at 24.4 mg/m^2^. The age of the LLINs was also an independent predictor of mosquito mortality (%) after 24 hours. The mortality decreased with increasing age of the LLIN. This model using segmented regression was able to predict 89.7% of the time if a LLIN will achieve the WHO 80% of mortality threshold for bio-efficacy using only two predictors: insecticide content measurements obtained from bromine x-ray fluorescence testing and the age of the LLINs.

**Figure 5.**
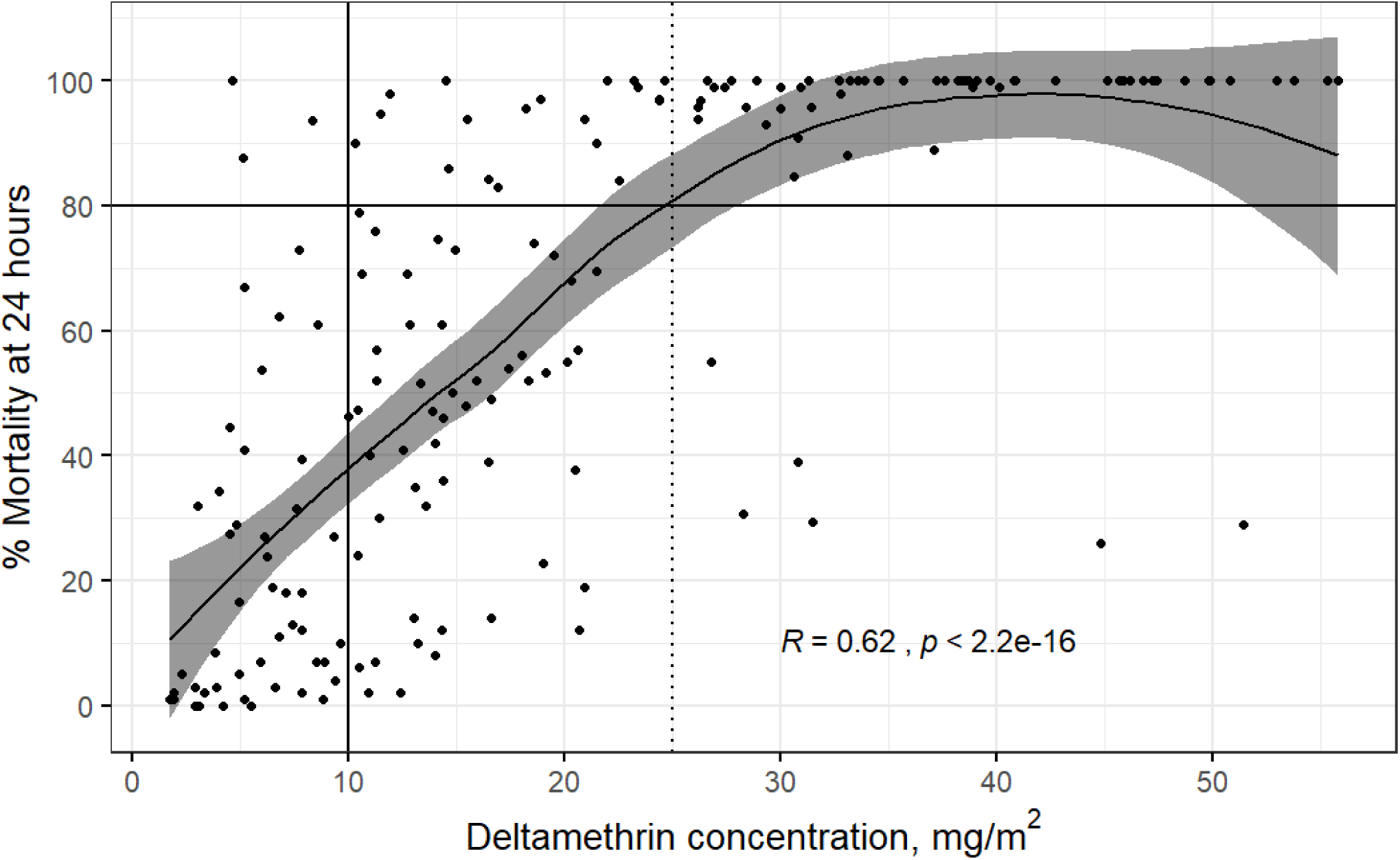
Locally weighted regression (LOESS) analysis between results of the cone bioassays measuring percent mortality at 24 hours and concentration of deltamethrin (mg/m^2^) as measured by X-ray fluorescence (XRF) analysis for all survey time-points. Each dot represents one long-lasting insecticide-treated bednet (LLIN) plotted based corresponding cone bioasays and XRF measurements. The gray area represents the 95% confidence interval. The solid vertical line represents the threshold of 10 mg/m^2^ and the dashed vertical line represents the threshold of 25 mg/m^2^. The black horizontal line represents the 80% mortality threshold.

### Overall proportion of LLINs providing adequate protection

The proportion of LLINs providing adequate protection was 38% (CI95: 33-44) at 24 months and 21% (CI95: 17-26) at 32 months. Three components contributed most to the observed decreases in adequate protection: 1) A decrease between the number of LLINs distributed and the proportion of LLINs still present in the household-which was particularly important at 32 months (drop of 34%); 2) A reduction between the proportion of LLINs present and used at least once to the proportion of LLINs present and used the night before of the survey (drop of 21%, 16% and 18% for 18, 24 and 32 months, respectively); and 3) A drop between the proportion of LLINs in serviceable condition and the proportion of LLINs in serviceable condition with a deltamethrin concentration of at least 10 mg/m^2^ (drop of 12% and 15% for 24 and 32 months, respectively) (Figure 6 and Table S4).

**Figure 6.**
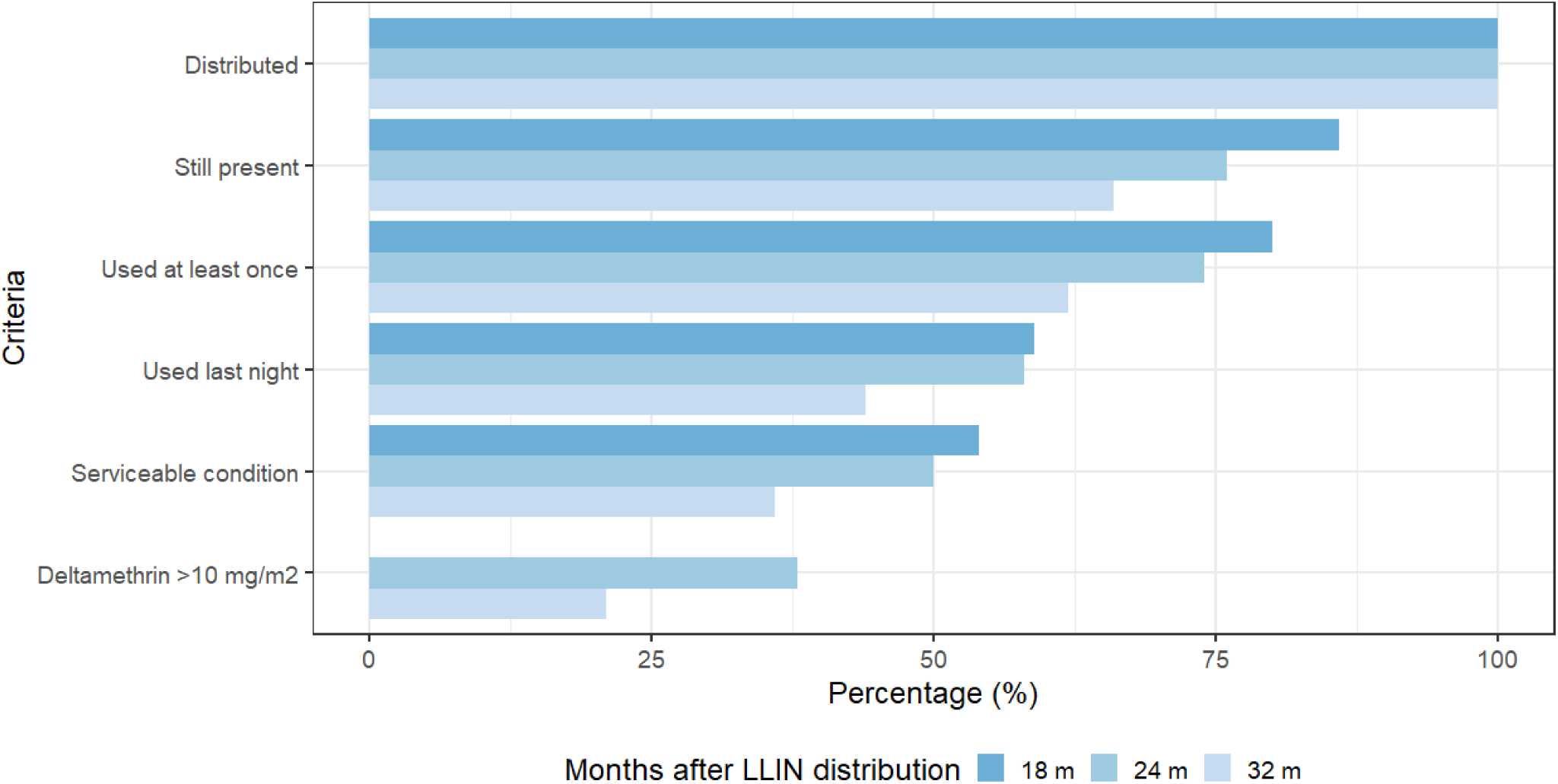
Estimate of effective protection of long-lasting insecticide-treated bednets (LLINs) in La Gomera, Escuintla. The proportion of LLINs that met each of the criteria listed on the left is shown in sequential descending order, with each value shown as a subset of the criteria above. If the LLIN was able to fulfill all criteria, it was considered a LLIN that provided adequate protection. We did not include the stage of deltamethrin above 10 mg/m^2^ as measured by XRF for the 18-month survey as we only analyzed a small number of LLINs with XRF at that time-point.

## DISCUSSION

In this prospective multiple cross-sectional study, we assessed the durability of PermaNet^®^ 2.0 LLINs distributed in a region of persistent malaria transmission in Guatemala by measuring their overall and functional survivorship, physical integrity, insecticide content and bio-efficacy.

The proportion of LLINs present in the household and in ‘serviceable’ condition was 69% after 32 months of routine use, with an extrapolated median survival time of 3.7 years. The median survival time reported in our study is higher than similar studies conducted in Africa that have also evaluated PermaNet^®^ 2.0 LLINs. In Tanzania the estimated survival was 2.5 years, in Zanzibar it was 3.1-3.3 years and in Zambia it was 2.5-3 years (Haji et al., 2020; Lorenz et al., 2019; Tan et al., 2016). In our study, the physical integrity of the LLINs—a key component for the functional survivorship—was high in each of the survey time-points (range: 84-94%), whereas other studies have reported lower values (e.g. 68% at 36 months) (Haji et al., 2020). These differences are further highlighted by the median total hole area, which in our study was only 34 cm^2^ at 32 months, whereas other studies evaluating PemaNet^®^ 2.0 LLINs reported values of 390 cm^2^ and 106 cm^2^ (Craig et al., 2015; Vanden Eng et al., 2015). In general, the size of the holes found on the LLINs in Guatemala was smaller, resulting in a greater number of LLINs categorized as being in ‘serviceable’ condition and a higher functional survivorship. The low proportion of new holes caused by thermal damage or animals as compared to other studies may also help explain these differences (Morgan et al., 2015; Mutuku et al., 2013). Of the LLINs containing holes, only 44% showed evidence of repair.

From a programmatic perspective, we also estimated the overall survivorship of the LLINs, which was 66% after 32 months. This was very similar to the functional survivorship, indicating that two thirds of the campaign LLINs were still present and functioning well in the surveyed households nearly 3 years after their distribution. However, reasons explaining the overall and functional survivorship were different in the 18- and 24-month surveys. At these time points, the main reason reported for not having a net present in the household was because it had been ‘given away to others’, not because of wear and tear. These findings are similar to previous reports (Gnanguenon, Azondekon, Oke-Agbo, Beach, & Akogbeto, 2014; Mansiangi et al., 2020; Obi et al., 2020), highlighting the real-life challenges faced by national malaria programs, which reach beyond the quality of the fabric of the nets. In a large study evaluating 14 household surveys from four African countries, 34% of the nets that were not present in the household had been given away, the majority to family members and within the first month after a distribution campaign (Koenker et al., 2014).

In terms of usage and handling, between 71-79% of the users reported to have used the LLINs the night preceding the survey, with almost a third of the nets observed not being in use as they were not hanging in the sleeping place. Previous studies that have evaluated the usage of LLINs in Latin America have shown a variety of results around LLIN usage. In Colombia, the usage was 51.1% after 8 months, whereas in Venezuela, it was over 90% after 6 months, highlighting intra-regional differences (Alvarado, García, Villarroel, & Rosas-Aguirre, 2011; Cabrera, Diaz, Pareja, & Santamaría, 2009).

We detected decreases in insecticide content over time, with a drop of 53% detected between unused nets (49.8 mg/m^2^) and nets at the 18-month survey point (23.2 mg/m^2^). These levels were lower than levels detected on PermaNet^®^ 2.0 in Ethiopia, where the mean concentration of deltamethrin was 44.1 mg/m^2^ after 14-20 months of use (Anshebo et al., 2014). In our study, the proportion of nets with deltamethrin levels ≥10 mg/m^2^ after 32 months was 65%, a stark contrast to the 95% observed in the aforementioned study. The values obtained by XRF to estimate total insecticide content showed similar trends to the bioassays, where just 52% of the LLINs at 32 months met the WHO bioassay threshold of bio-efficacy. The frequent washing of the LLINs and the high proportion that were dried under direct sunlight are some of the factors that could have contributed to this decline (Agossa et al., 2014; Atieli, Munga, Ofulla, & Vulule, 2010). As other studies have detected, we also found that a measurement of ≥25 mg/m^2^ by XRF was more likely to indicate a LLIN with a mortality at 24 hrs. of ≥80%, suggesting that this threshold is more closely associated with optimal bio-efficacy than 10 mg/m^2^ (Anshebo et al., 2014).

When we integrated the results of survivorship, physical integrity, usage and insecticide content, the estimated that the level of protection provided by the LLINs was sub-optimal. The overall proportion of LLINs providing adequate protection was only 38% after 24 months and dropped further to 21% at 32 months. National malaria data showed that the number of cases in Escuintla remained constant during the period of 2012-2015, suggesting that the LLINs did not provide sufficient protection.

Several lessons can be gathered from our findings. First, we suggest that three key factors contributed to the sub-optimal level of protection: the loss of LLINs still present in the household, the degree of usage of the LLINs, and the low percentage of LLINs with adequate insecticide content. Given that these factors are potentially compromising the protective effectiveness of the LLINs more than the durability of the fabric, activities focusing on community engagement and the inclusion of local civil society around LLIN use could be particularly impactful. Second, our findings suggest that personal redistribution of nets is an important trend in this region, similar as in Africa. By giving nets away to friends and family, communities may be compensating for inadequate campaign planning. National malaria programs may consider including local leadership and shift toward a more decentralized approach to improve the design of LLIN campaign distributions (Gosling et al., 2020). Third, we were able to show that in areas with limited resources to carry out the WHO cone bioassay, the measurement of insecticide content by bromine x-ray fluorescence might be an accurate alternative methodology to estimate the bio-efficacy of LLINs.

A major strength of our study is that we concurrently measured survivorship, physical integrity, insecticide content and bio-efficacy. There are few data on these indicators reported from Latin America, and our study provides one of the most comprehensive evaluations to date for the Americas. In addition, by following standard WHO guidelines, our results can be compared to outcomes arising from similar studies conducted elsewhere, while at the same time including variables that were customized to the local context. The use of GIS in the 24- and 32-month surveys provided us with a more accurate identification of the households in each community, and our randomization process allowed us to limit bias associated with the selection of the surveyed households. However, our study also had several limitations. First, we were not able to conduct a household survey at the 6- or 12-month time-points. Second, we had to exclude four out of the twelve selected communities in the 32-month survey, which could introduce selection bias. Third, the answers for the questions related to usage, care and handling of the LLINs are prone to recall and social desirability bias (Althubaiti, 2016).

## CONCLUSION

The lifetime of PermaNet^®^ 2.0 is expected to be approximately 3 years. In our study, after 2 years, over two thirds of the LLINs did not contain the optimal deltamethrin concentration of 25 mg/m^2^ and 32% of them did not meet the WHO criteria for bio-efficacy. Even though the nets were in good physical condition at 32 months, their diminished bio-efficacy and reported lack of consistent use compromised the protection provided to the users. The widespread provision of LLINs is a cornerstone of global efforts for the control and potential elimination of malaria, which is now a near-term goal in several Latin American countries (World Health Organization, 2018). We recommend that future evaluations of the durability of LLINs should always include measurements of survivorship, usage and handling of the LLIN, physical integrity, and insecticidal activity (insecticide content and/or bio-efficacy) in order to estimate the protective effect of the nets in a community. The engagement of the National Malaria Program with the community is critical to the success of future LLIN distributions in Guatemala.

## Data Availability

The data that support the findings of this study are available on request from the senior author, NP. The data are not publicly available due to containing information that could compromise the privacy of participants.

## LIST OF ABBREVIATIONS

GIS: geographic information system
GIS: geographic information system
GPS: Global positioning system
IQR: interquartile range
ITN: Insecticide-treated bednet
LLIN: long-lasting insecticide-treated bednet
LOESS: Locally Weighted Scatterplot Smoothing
MoH: Ministry of Health
PDA: personal digital assistant
THSA: total hole surface area
tm: median survival time
WHO: World Health Organization
XRF: x-ray fluorescence

## DECLARATIONS

### Ethics approval and consent to participate

Oral informed consent was obtained from all participants prior to study inclusion. This study was approved by the Ethics Committee of the Center for Health Studies at Universidad del Valle de Guatemala (Approval Number: 081-06-2013); CDC investigators were not considered to be engaged in human subjects research.

### Consent for publication

Not applicable

### Author’s contributions

Conception and design: NP, AL, SC, ZM, JVE, SS, MEC; survey data collection, supervision and coordination: SC, NP, MEC, JCL, JGJ, insecticide content and bio-efficacy assays: SR, LV, SS; data analysis: MEC, JVE, HV, SR, draft of manuscript: MEC, SR, review and edit of the manuscript: MEC, SR, JCL, JGJ, ZM, SC, SS, LV, JVE, HW, AL, NP. All authors approved the final version of the manuscript.

## Acknowledgements

We thank the invaluable contribution of the survey team from the National Malaria Sub-program, Vector-borne Diseases Program and from Universidad del Valle de Guatemala, particularly Rodrigo Flores, Pedro Peralta, and Alfonso Salam. We thank Gerard Lopez for his help in the design of the survey and installation in the PDAs. We also thank all the participants in the survey and all the help provided by the community leaders at La Gomera, Escuintla. We thank Vestergaard for providing us with LLINs to replace the ones we used for the laboratory analyses. We are grateful to Barbara Marston and William Hawley for their critical suggestions and for their careful reading of the manuscript.

## Disclaimer

The findings and conclusions in this paper are those of the authors and do not necessarily represent the official position of the Centers for Disease Control and Prevention.

## Tables Legends

Table 1. Demographic characteristics of the surveyed households and head household at the three time-points.

Table 2. Long-lasting insecticide-treated bednets (LLINs) use and handling as reported by householders at each survey time-point.

## FIGURE LEGENDS

## SUPPLEMENTARY MATERIAL

### Table Legends

**Table S1.**
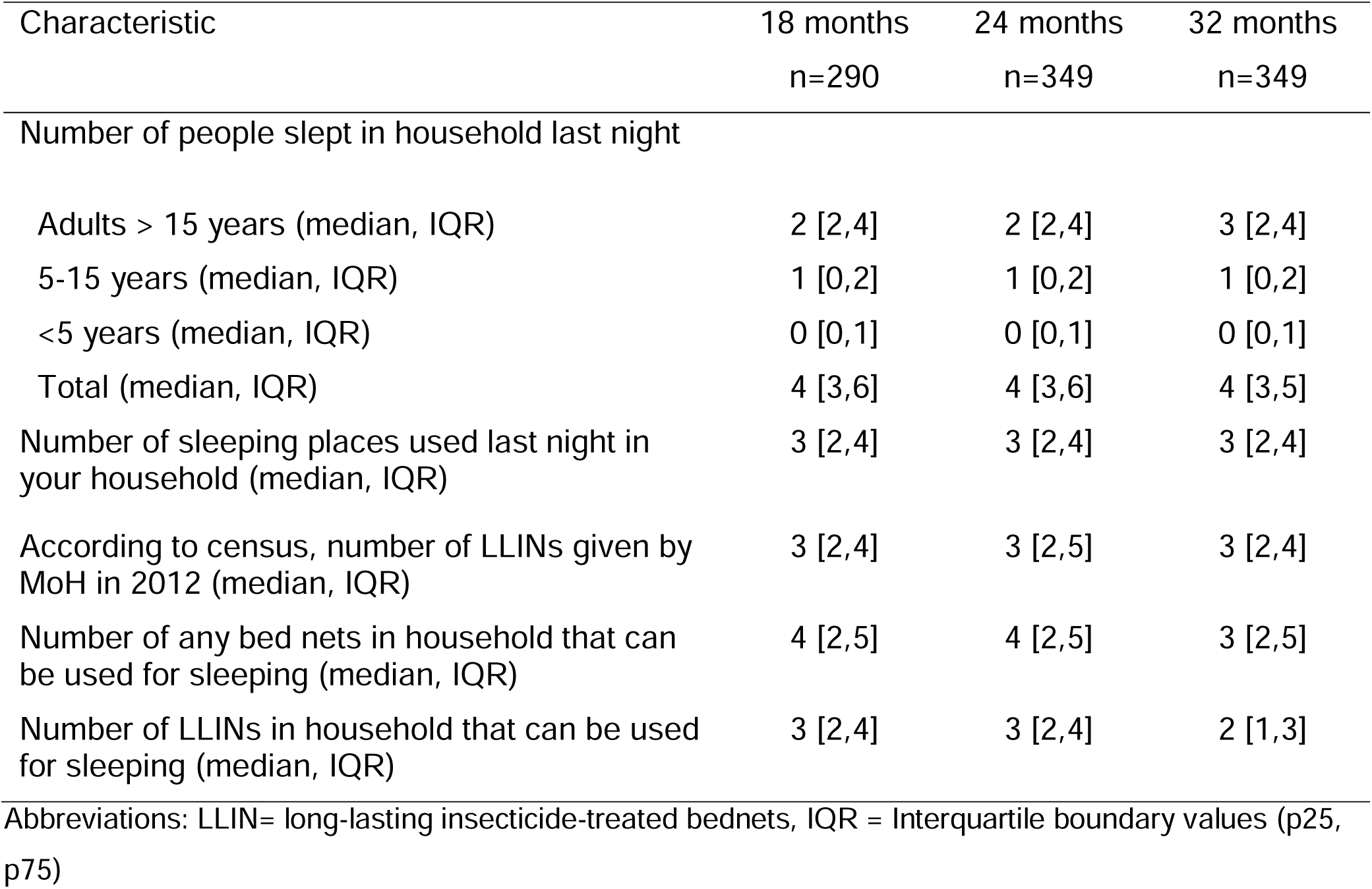
Additional descriptive characteristics of the households that were enrolled in the surveys at the three time-points.

**Table S2.**
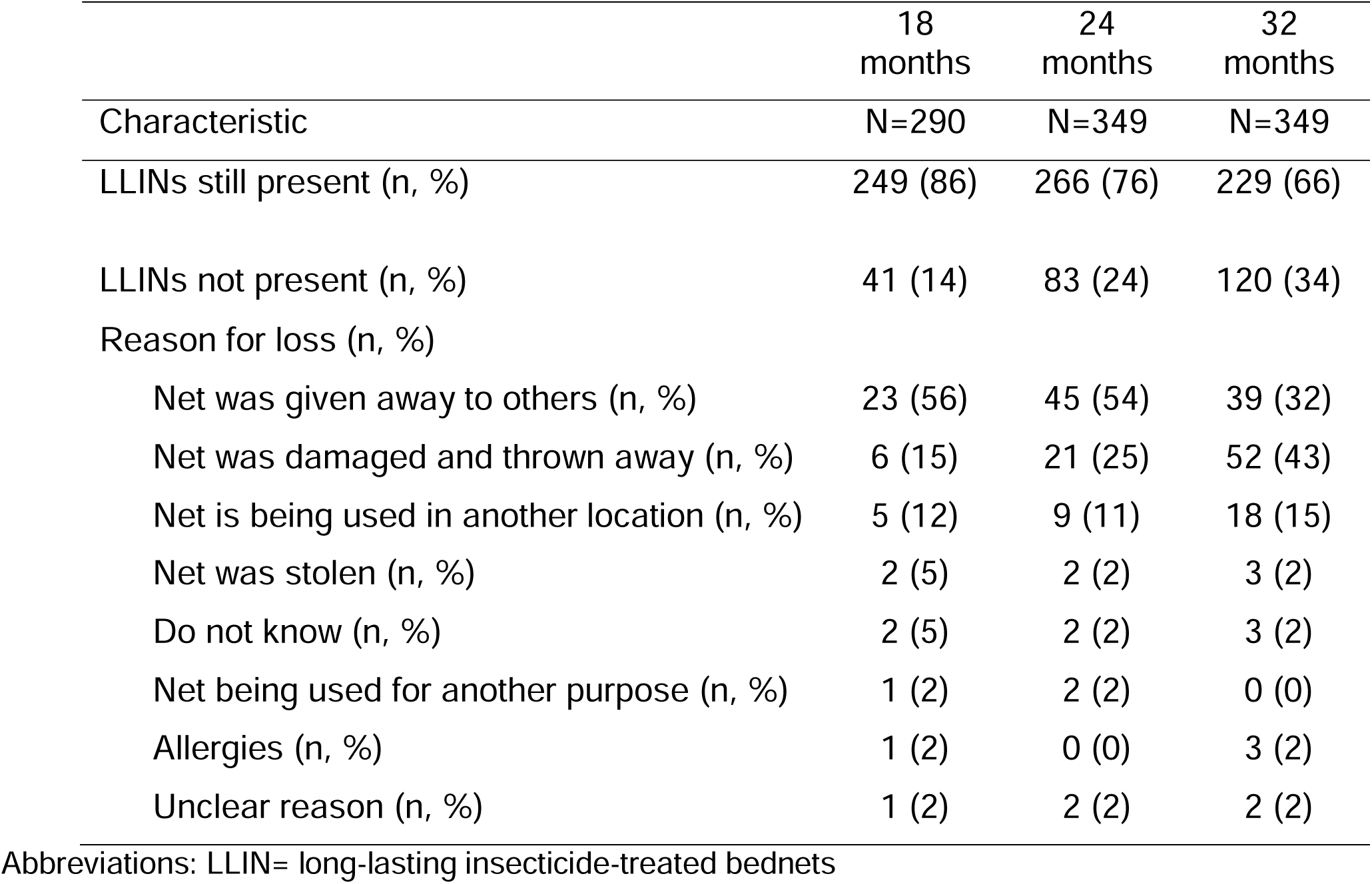
Reasons for LLIN loss.

**Table S3.** Association of deltamethrin (mg/m^2^) content level measured by X-ray fluorescence (XRF) and age of the long-lasting insecticide-treated bednets (LLINs) in predicting the percentage of mosquito mortality within 24 hrs.

| Exposure variable (1st segment) | Estimate (se) | p-value |
| --- | --- | --- |
| Deltamethrin (mg/m<sup>2</sup>) in first segment | 2.8 (0.3) | <0.001 |
| 18 months | 27.1 (5.6) | <0.001 |
| 24 months | -18.8 (4.3) | <0.001 |
| 32 months | -24.9 (4.3) | <0.001 |

Association of Deltamethrin (mg/m<sup>2</sup>) in the 2nd segment was 1.1 (se = 0.5, CI95: 0.2, 2.0)

**Table S4.** Estimate of effective protection of long-lasting insecticide-treated bednets (LLINs) in La Gomera, Escuintla. Results are shown for a) the overall study population, b) Data excluding LLINs with incomplete hole section and c) Data excluding LLINs with incomplete hole section and with no X-ray fluorescence (XRF) analysis. Results in bold are the ones presented in Figure 6.

| Criterion | 18 months | 24 months | 32 months |
| --- | --- | --- | --- |
| Overall data |  |  |  |
| Evaluated<sup>±</sup> (n, %) | **290 (100)** | **349 (100)** | **349 (100)** |
| Still present (n, %) | **249 (86)** | **266 (76)** | **229 (66)** |
| Used at least once (n, %) | **231 (80)** | **257 (74)** | **217 (62)** |
| Used last night (n, %) | **171 (59)** | **203 (58)** | **154 (44)** |
| Data excluding LLINs with incomplete hole section |  |  |  |
| Evaluated<sup>π</sup> (n, %) | 281 (100) | 348 (100) | 346 (100) |
| Still present (n, %) | 240 (85) | 266 (76) | 226 (65) |
| Used at least once (n, %) | 222 (79) | 256 (74) | 214 (62) |
| Used last night (n, %) | 164 (58) | 203 (58) | 152 (44) |
| Serviceable condition (based on total hole area) (n, %) | **151 (54)** | **174 (50)** | **126 (36)** |
| Data excluding LLINs with incomplete hole section and with no XRF analysis |  |  |  |
| Evaluated<sup>π π</sup> (n, %) | 91 (100) | 346 (100) | 335 (100) |
| Still present (n, %) | NE | 263 (76) | 215 (64) |
| Used at least once (n, %) | NE | 254 (73) | 204 (61) |
| Used last night (n, %) | NE | 201 (58) | 145 (43) |
| Serviceable condition (based on total hole area) (n, %) | NE | 172 (50) | 121 (36) |
| Over 10 mg/m<sup>2</sup> (based on XRF analysis) (n, %) | NE | **133 (38)** | **72 (21)** |

Abbreviations: LLIN= long-lasting insecticide-treated bednets, XRF = X-ray fluorescence
± This 'n' serves as the denominator for the overall data analysis.
π This 'n' serves as the denominator for the data excluding LLINs with incomplete hole section
π π This 'n' serves as the denominator for the data excluding LLINs with incomplete hole section and with no XRF measurement.
NE =Not estimated because only a small proportion of LLINs were analyzed by XRF analysis, so this measurement is likely to be biased.

## SUPPLEMENTARY MATERIAL

### Figure Legends

**Figure S1.**
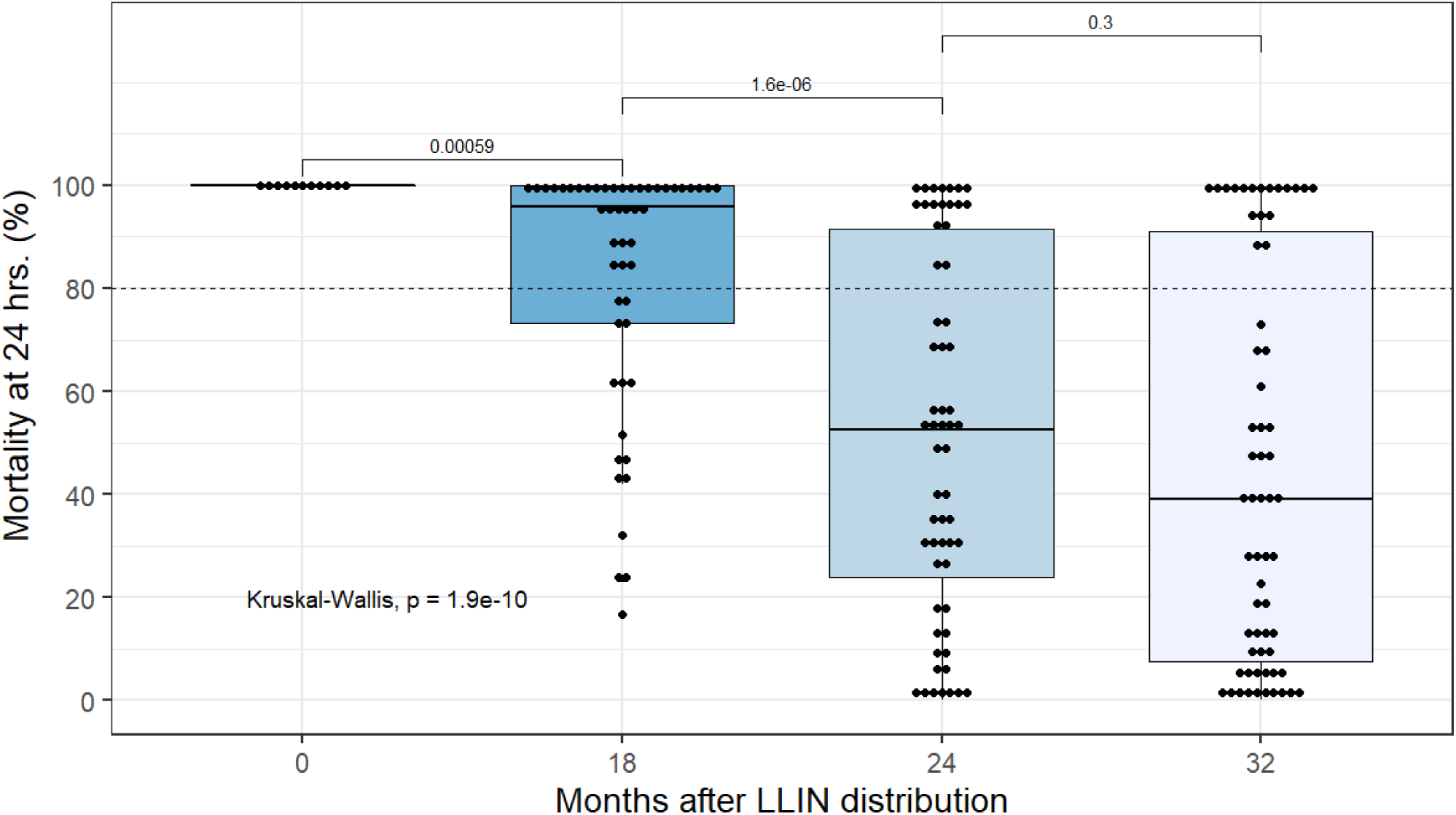
Mortality at 24 hours of *An. albimanus* (Sanarate strain) mosquitoes exposed in cone bioassays to long-lasting insecticide-treated bednets (LLINs) at the surveys time-points. The time-point of 0 represents values on unused LLINs that were from the same batch as those distributed. Each box represents the interquartile distance, center line in each box indicates the median. The black dots indicate each of the values of a LLIN. The dashed horizontal line represents the 80% mortality threshold.

**Figure S2.**
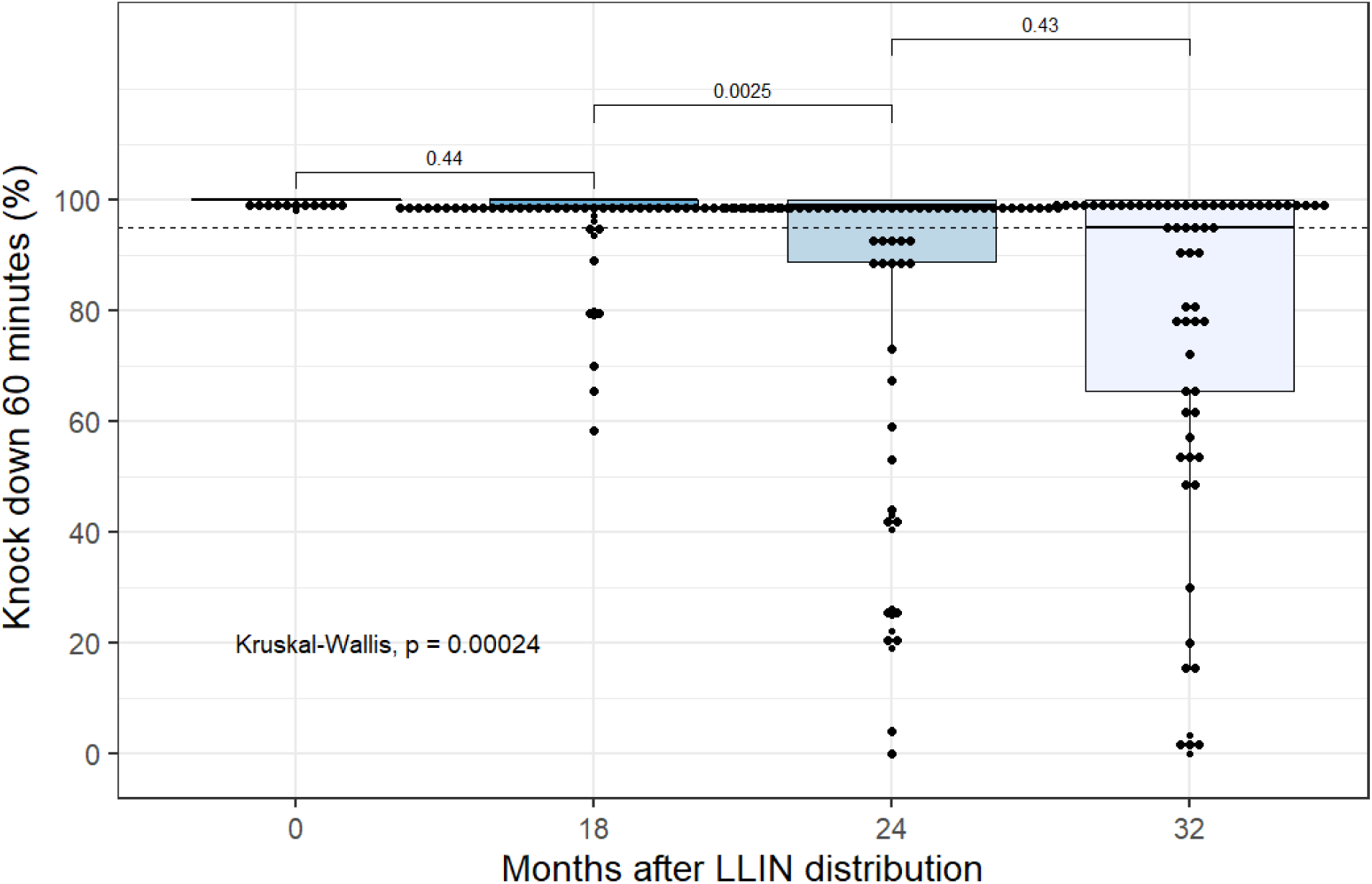
Knockdown after a 60-min (KD60) exposure of *An. albimanus* (Sanarate strain) mosquitoes exposed in cone bioassays to long-lasting insecticide-treated bednets (LLINs) at the surveys time-points. The time-point of 0 represents values on unused LLINs that were from the same batch as those distributed. Each box represents the interquartile distance, center line in each box indicates the median. The black dots indicate each of the values of a LLIN. The shaded horizontal line represents the 95% KD60 threshold.

**Figure S3.**
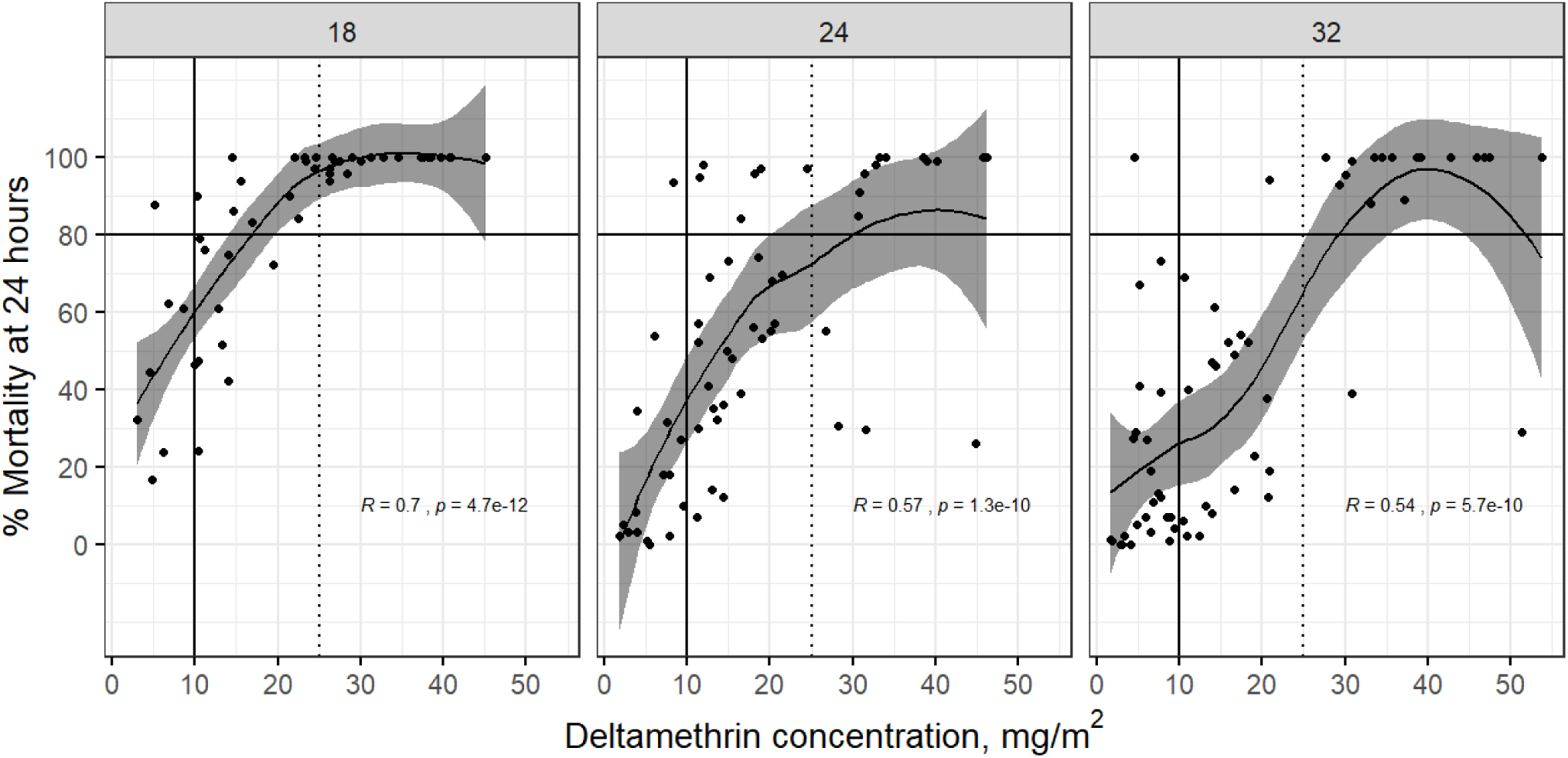
Locally weighted regression (LOESS) analysis between results of the cone bioassays measuring percent mortality at 24 hours and concentration of deltamethrin (mg/m^2^) as measured by X-ray fluorescence (XRF), stratified by surveys time-points (18 months, 24 months an 32 months). Each dot represents one long-lasting insecticide-treated bednets (LLINs) with their corresponding cone bioasays and XRF measurements. The gray area represents 95% confidence interval. The solid vertical line represents the threshold of 10 mg/m^2^ and the dashed vertical line represents the threshold of 25 mg/m^2^. The black horizontal line represents the 80% mortality threshold.

## APPENDIX FILE

### Methodology to assess if insecticide concent measurement obtained by XRF could serve as a predictor to estimate the mortality at 24 hours

- We developed a predictive model using piecewise (segmented) regression to accommodate the change in the magnitude and direction of the linear relationship between the outcome and exposure variables observed in our exploratory analysis.

We use selected responses from the surveys as exposure variables:

− Months after distribution of LLIN
− Highest level of education of the head of the household
− Net still present in the household
− Net used the night before of the survey to sleep under
− Frequency of the net used in the week before of the survey to sleep under
− Adult (>15 years) slept under the net the night before of the survey
− Children (5-15 years) slept under the net the night before of the survey
− Children (<15 years) slept under the net the night before of the survey
− Net has been washed
− Last time the net was washed
− Type of soap used to wash the net
− Net was soaked when washing
− Length of time a net was soaked
− Net was scrubbed when washing
− Location were net was dried
− Presence of open flame were the net was located
− Total area of holes of LLIN
− Condition of LLIN based on holes area (Good, damaged or too torn)
− Deltamethin content (mg/m^2^) measured by XRF
− Outcome variable: % Mortality at 24 hours
− The model was first trained using 80% of the data and validated in the remaining 20%. We used random sampling to separate the data as training and test data. This process was repeated 1000 times. To assess the accuracy of the fitted model, a confusion matrix was built to calculate the proportion of correct classification when the model was applied to the test data

